# A Human Skin Model for Assessing the Arboviral Infections

**DOI:** 10.1101/2022.02.17.22271132

**Authors:** Allen T. Esterly, Megan G. Lloyd, Prashant Upadhyaya, Jennifer F. Moffat, Saravanan Thangamani

## Abstract

Arboviruses such as flaviviruses and alphaviruses cause a significant human healthcare burden on the global scale. Transmission of these viruses occurs during human blood feeding at the mosquito-skin interface. Not only do pathogen immune evasion strategies influence the initial infection and replication of pathogens delivered, but arthropod salivary factors also influence transmission foci. In-vitro cell cultures do not provide an adequate environment to study complex interactions between viral, mosquito, and host factors. To address this need for a whole tissue system, we describe a proof-of-concept model for arbovirus infection using adult human skin ex vivo with Zika virus (flavivirus) and Mayaro virus (alphavirus). Replication of these viruses in human skin was observed up to 4 days post-infection. Egressed viruses could be detected in the culture media as well. Antiviral and pro-inflammatory genes, including chemoattractant chemokines, were expressed in infected tissue. Immunohistochemical analysis showed the presence of virus in the skin tissue at 4 days post-infection. This model will be useful to further investigate: 1) the immediate molecular mechanisms of arbovirus infection in human skin, and 2) the influence of arthropod salivary molecules during initial infection of arboviruses in a more physiologically relevant system.

## Introduction

Arboviruses inflict a substantial global health burden on the human population. The re-emergence of Zika virus (ZIKV) in 2014-2015 in Brazil is evidence that previously identified pathogens can re-emerge to become significant health burdens on susceptible populations (Weaver et al., 2016). Other arboviruses, such as Usutu virus (USUV), Spondweni virus (SPOV), and Mayaro virus (MAYV), continue to emerge (Pierson & Diamond, 2020). MAYV is a New World arthritogenic alphavirus with current autochthonous cycles of transmission in Central and South America (Diagne et al., 2020). Research on emerging arboviruses is needed to better understand transmission cycles that threaten human health.

Mosquitoes inject saliva made up of a complex mixture containing pathogens, bioactive proteins, and small RNAs to facilitate blood-feeding and pathogen transmission (Arcà & Ribeiro, 2018; Maharaj et al., 2015). These bioactive molecules regulate hemostasis and modulate mammalian immune responses at the mosquito-host-pathogen interface (Huang et al., 2019; Pingen et al., 2017). The transmission interface has been investigated using in-vitro tissue culture or animal infection models, but these systems do not fully represent the human skin tissue environment. Therefore, this study intends to model arbovirus infection in human skin ex vivo to investigate the immediate immunological events during transmission.

Since the majority of mosquito-borne arboviruses transmitted to humans are from the genus Flavivirus or Alphavirus, we used an established flavivirus of medical relevance (ZIKV) and an emerging alphavirus (MAYV). MAYV and ZIKV infect human skin and replicate up to 4 days post-infection, shedding from the skin to culture media. Chemoattractant chemokines, along with pro-inflammatory and antiviral genes, were expressed in both MAYV and ZIKV infected skin as expected. In addition, immunohistochemical analysis revealed staining positive for the virus in skin 4 days post-infection in both virus-infected groups. This model will be further utilized to determine how mosquito salivary factors influence viral replication kinetics or pro-inflammatory and chemoattractant molecule expression during infection.

## Materials & Methods

### Ethics Statement

All experiments involving de-identified human specimens were conducted in a biological safety level 2 (BSL-2) laboratory in accordance with a protocol approved by the SUNY Upstate Medical University Institutional Review Board (IRB).

### Culturing of Cells and Viruses

Vero, Vero E6, and C6/36 cells were obtained from ATCC and passaged in DMEM with 10% heat-inactivated fetal bovine serum (HI-FBS) and 1% penicillin-streptomycin at 37°C + 5% CO2 (Vero & Vero E6) or in Leibovitz L-15 media with 10% HI-FBS and 1% penicillin-streptomycin at 28°C (C6/36) (Ammerman et al., 2008). Zika virus (Mex I44) was obtained from the World Reference Center for Emerging Viruses and Arboviruses at the University of Texas Medical Branch in Galveston, Texas. Mayaro virus (Uruma) was obtained from the Centers for Disease Control, Fort Collins, Colorado. Viruses were cultured in Vero cells (ATCC CCL-81) for 4 passages and then passaged once in C3/36 cells as referenced (Coelho et al., 2017; Svenson et al., 2018). In addition, virus titers were obtained via focus-forming assay in Vero E6 cells as previously described (Rossi et al., 2012).

### Preparation and Infection of human skin

De-identified adult human skin was donated by fully informed patients undergoing reduction mammoplasty at the Upstate Medical University Hospital (Syracuse, NY). All skin specimens were collected within approximately 2 h post-surgery. Underlying adipose was grossly dissected, and each specimen was washed in PBS and incubated in skin culture media (RPMI 1640 media containing 10% HI-FBS, 1% penicillin-streptomycin, and 0.25ug/mL of Amphotericin B)(Lloyd et al., 2020). The epidermis and upper dermal layers were removed using a skin grafting knife, cut into 1-cm^2^ biological samples, then placed in Netwells^®^ (Corning, 15-mm x 500 µm mesh size) suspending the skin at a liquid-air interface. Netwells were placed in 12-well tissue culture plates containing 1 mL skin culture media. The skin was cultured ex vivo for 4 days in skin culture media at 33°C with 5% CO2 as previously described (Lloyd et al., 2020). Next, the skin was infected intradermally using a 28-gauge insulin syringe with 25 µL (±2 µL) of uninfected skin culture media (mock), ZIKV, or MAYV adjusted to deliver 10^3^ FFU. Skin samples were processed as outlined below at 2- and 4-days post-infection. Each experimental group contained multiple biological replicates (n=4) and independent infections were repeated using skin specimens from 3 donors.

### RNA Isolation, qRT-PCR, and Gene Expression Analysis

At days 2 and 4 post-infection, skin samples were cut in half using a scalpel and placed in either 10% neutral buffered formalin or OCT media for histological analysis or in TRIzol reagent for RNA isolation. RNA was extracted from the skin and skin culture media samples using TRIzol and TRIzol LS reagents, respectively, combined with Qiagen RNeasy mini kits to inactivate the virus and obtain high yields of RNA(Heinze et al., 2012; Hermance et al., 2020). RNA purity and concentration were verified on a Denovix DS-11+ spectrophotometer.

Absolute quantification of viral loads was performed using quantitative reverse transcriptase PCR (qRT-PCR) using a BioRad CFX96. Viral genomes were amplified with the BioRad One-Step Universal Probes kit using primers: ZIKV 1086, ZIKV 1162c, and probe ZIKV 1107-FAM or MAYV Forward (5’-AAGCTCTTCCTCTGCATTGC-3’, MAYV Reverse 2 (5’-TGCTGGAAATGCTCTTTGTA-3’), and MAYV Probe (5’-GCCGAGAGCCCGTTTTTAAAATCAC-3’) previously established (Lanciotti et al., 2008; Waggoner et al., 2018). The MAYV primer-probe combination was slightly modified to utilize a Cy5/Iowa Black RQ fluorophore/quencher combination from Integrated DNA Technologies (Coralville, Iowa). RNA isolated from an aliquot of both ZIKV and MAYV, from the exact passage used for infection, was serially diluted, and used to design a standard curve to quantify viral loads in tissue and supernatant.

cDNA conversion of tissue RNA was performed according to the manufacturer’s specification using the BioRad iScript cDNA synthesis kit using approximately 1µg of RNA. cDNA was diluted 5-fold, and gene expression analysis was performed using BioRad iTaq Universal SYBR Green Supermix according to the manufacturer’s specifications. Briefly, 1 µg of RNA/sample was used for the reverse transcriptase reaction. cDNA was diluted in RNase-free water at a 1:5 ratio, and a standard volume was used for each downstream qPCR reaction. Primers used for gene expression analysis are listed in Table 1. Expression was normalized using a eukaryotic ribosomal 18s as a ‘housekeeping’ gene with data shown as a ratio of gene target fold change in infected tissue relative to mock-infected tissue using the Livak (2^-ΔΔCq^) method (Livak & Schmittgen, 2001).

**Table 1.**
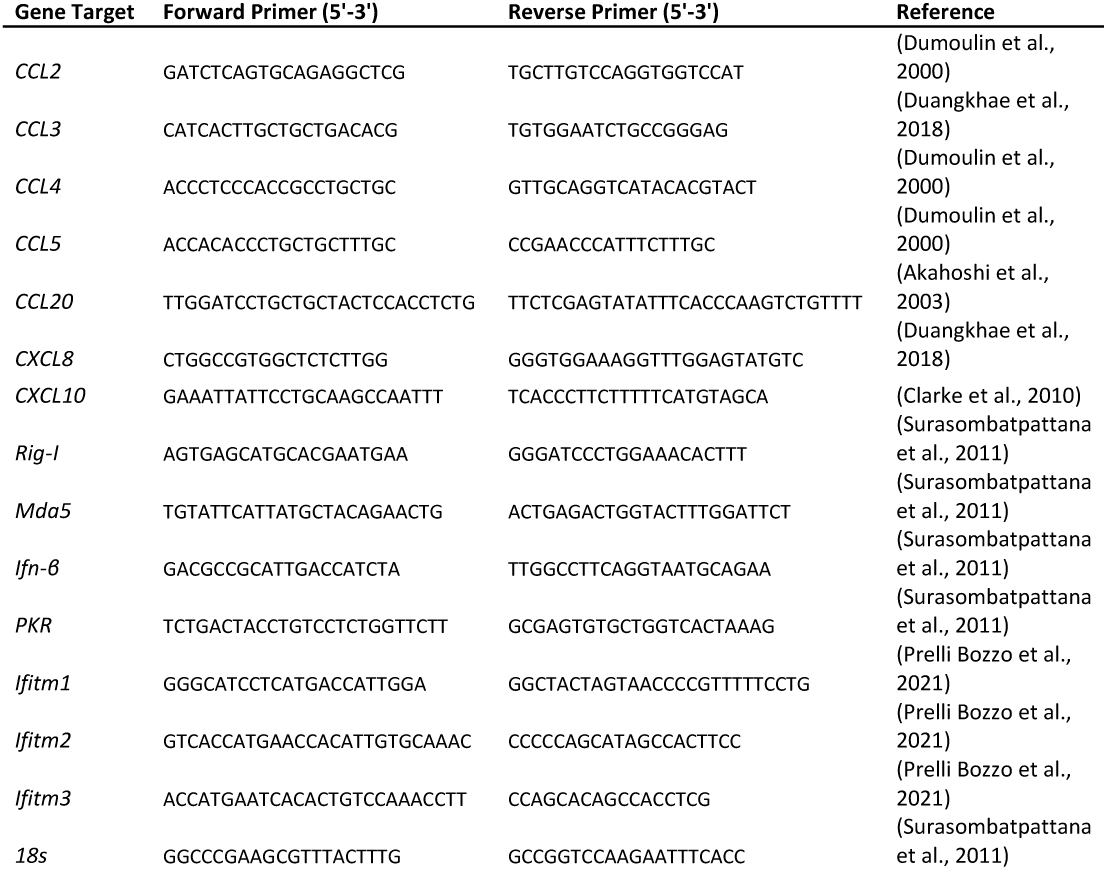
qPCR primers used for gene expression analysis.

### Histology and Immunohistochemistry

Tissues fixed in formalin were paraffin-embedded and cut in 5 µm thick sections to be placed on glass slides (Hermance et al., 2016). Samples were deparaffinized and dehydrated in xylene and graded ethanol washes. Hematoxylin and Eosin (H&E) staining was performed according to the manufacturer’s specifications (Vector Laboratories H-3502). Immunohistochemical detection of ZIKV and MAYV was performed using 5 µm frozen sections of tissues snap-frozen in OCT media immediately after the indicated endpoint. Briefly, tissue sections were fixed in methanol then washed in TBS+0.05% Tween20. Wash buffer was applied between all blocking and antibody incubation steps. Sections were treated with BLOXALL solution (Vector Labs, Burlingame, CA) to block endogenous peroxidases and alkaline phosphatases. Primary and secondary antibodies (murine immune ascites fluid generated against ZIKV or MAYV; goat anti-mouse HRP conjugate, respectively) were incubated for 1 h. Detection was performed with Vector Labs ImmPACT AMEC Red substrate, peroxidase kit. Cover slides were mounted using Vector Labs H5000 permanent mounting medium. Mock infected tissues were sectioned and treated, in parallel, with the same antibody incubation and wash steps as listed above. All images were taken using a Leica DM2500 LED/DMC5400 brightfield microscope and Leica Application Suite X (LAS-X) software.

### Data Analysis

Statistical significance of viral growth kinetics was performed using 2-way ANOVA with Šidák’s multiple comparisons test in GraphPad Prism 9.1.2. Statistical significance of gene expression data was analyzed by an F-test to determine variance, followed by a student’s t-test to assess the increase of the target gene from 2-4-days post-infection. Welch’s correction was applied when the F-test hypothesis was failed to be rejected.

## Data availability Statement

Data generated in study are paresend within this manuscript.

## Results and Discussion

### Viability of human skin explant

This study utilized human skin donated from elective breast reduction surgeries and cultured it ex vivo as previously described (Lloyd et al., 2020). Human skin was viable when cultured ex vivo for at least 4 days. Tissue sections stained with H&E showed minor histopathological changes after 4 days in culture. In addition, some papillary dermal edema and minimal lymphocytic infiltrate in the superficial dermis was observed; however, no major pathological changes, such as acanthosis or psoriasiform hyperplasia, were apparent (Figure 1).

**Figure 1.**
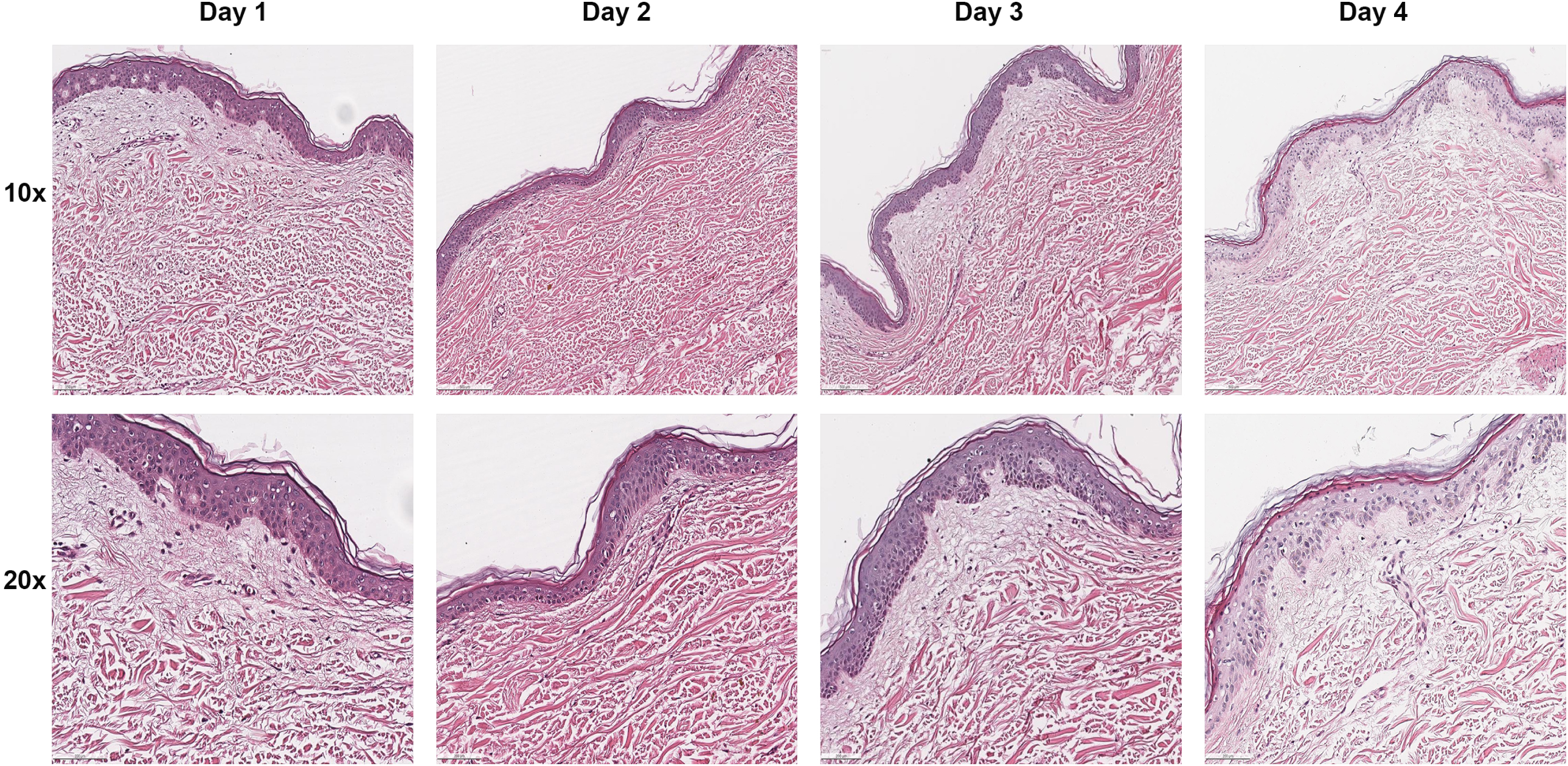
Viability of human skin ex vivo. Skin cultured ex vivo was harvested each day for up to 4 days. Formalin fixed, paraffin-embedded tissue sections (5 µm) were placed on glass slides for H&E staining. Pathological interpretation was provided by a board-certified pathologist (HISTOWIZ, Brooklyn, NY). Sections are representative of multiple biological samples from multiple skin specimens sectioned and stained using the same methodology.

### MAYV and ZIKV replication in human skin

To determine the susceptibility of human skin to arbovirus infection, skin pieces were intradermally inoculated with 1000 FFU of MAYV or ZIKV, a physiologically relevant dose delivered from a mosquito (Castanha et al., 2020; Styer et al., 2007). Viral RNA in the skin tissue was measured by qRT-PCR and increased for both MAYV and ZIKV during the 4-day incubation period (Figure 2). We also found that viral RNA in the skin culture media increased for both infections (Figure 2). Furthermore, in cryo-sectioned skin, immunohistochemical detection of MAYV and ZIKV showed widespread infected cells after 4 days (Figure 3), consistent with viral RNA detection in the homogenized tissue (Figure 2). Therefore, cultured human skin was viable and susceptible to arbovirus infection for up to 4 days.

**Figure 2.**
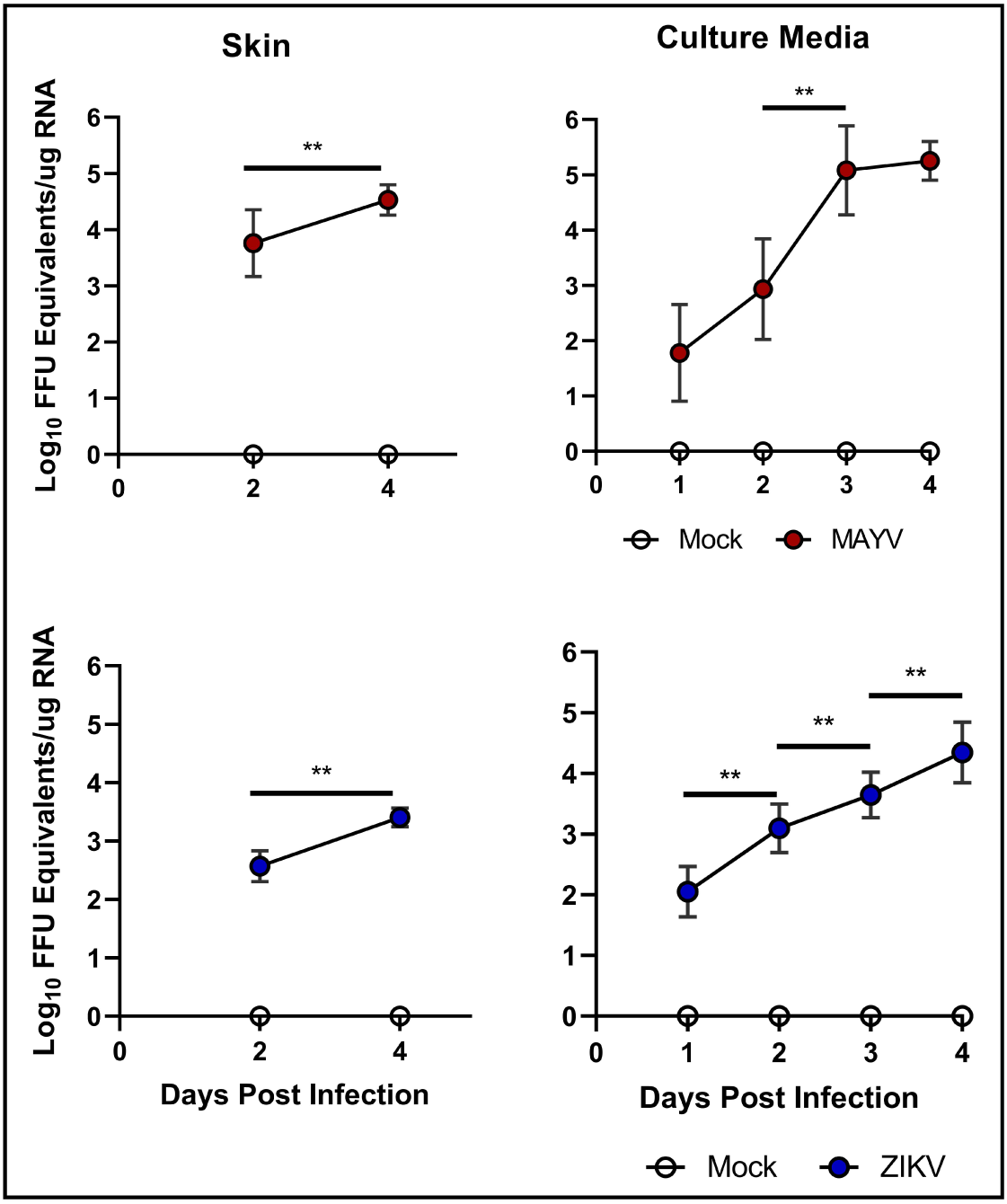
Skin is susceptible to arboviruses. Skin was infected intradermally with either MAYV (red) or ZIKV (blue). Data points are mean values with error bars indicating standard deviations. Data represents repeated independent experiments using n=4 biological replicates per group across multiple donated tissues. A two-way ANOVA was used to determine statistical difference with Šidák’s multiple comparisons \*\**p*<0.001.

**Figure 3.**
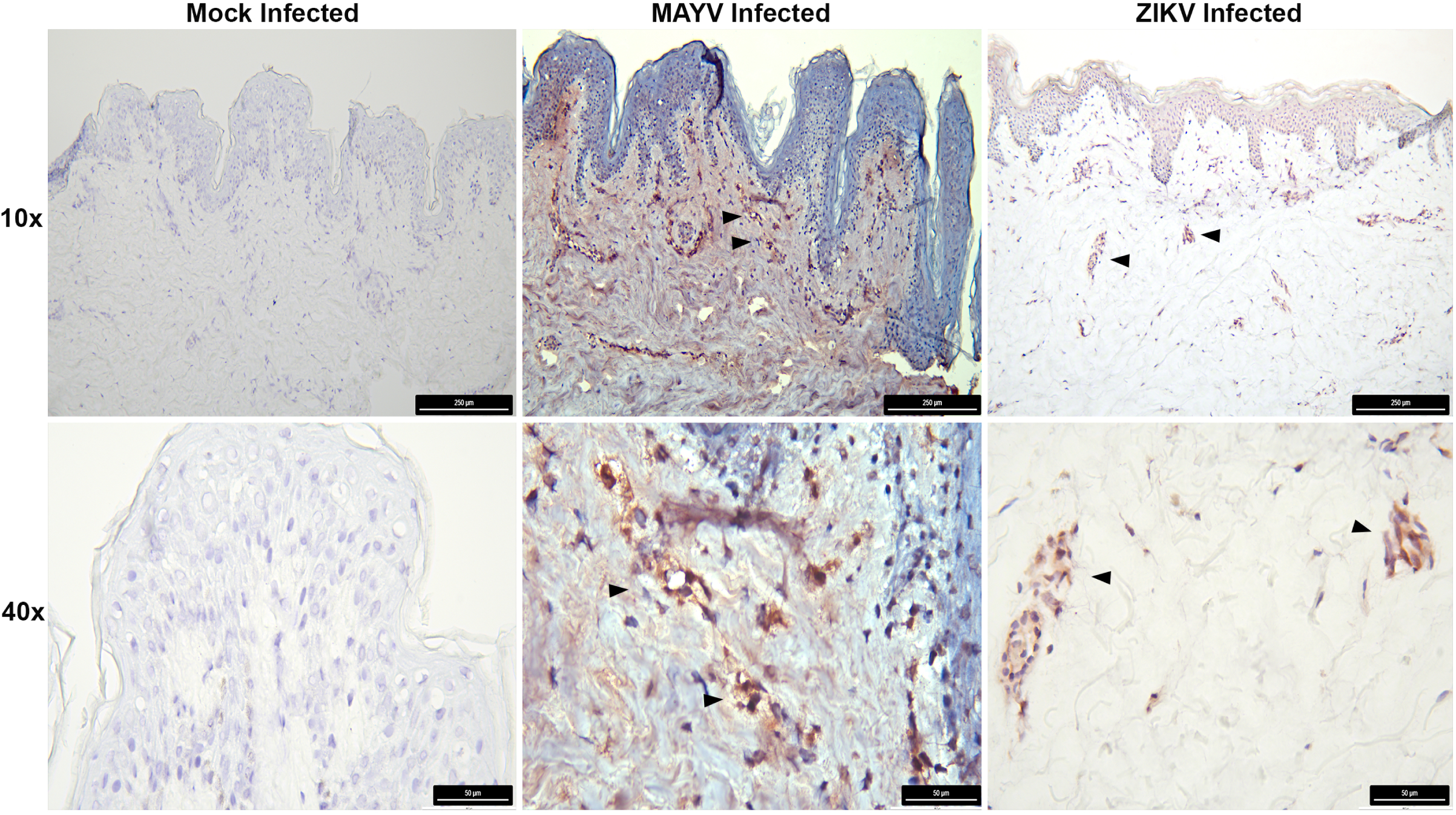
Expression of viral antigen in human skin. Immunohistochemical detection of MAYV antigen (middle column) and ZIKV antigen (right column) in cryo-sectioned skin post-infection. Arrows indicate antigen detection at 10x and 40x magnification. Mock tissues were treated with either MAYV or ZIKV antibodies in parallel to respective infected tissues. Skin sections are representative of multiple biological replicates from independent experiments with consistent detection of viral antigen.

Human skin processed similarly and infected with DENV-2 showed the presence of negative-strand RNA at 48h and 72h post-infection, indicative of viral replication (Limon-Flores et al., 2005). ZIKV infects several skin cell populations. Infection in primary cultures of epidermal keratinocytes and dermal fibroblasts show increasing viral RNA loads and infectious titer over time (Hamel et al., 2015). Dermal dendritic and Langerhans cells, present in the dermis and epidermis, respectively (Pasparakis et al., 2014), are also permissive to ZIKV infection when derived from PBMCs (Hamel et al., 2015). Our results showed increased replication of ZIKV in human skin explants over time which parallel previous findings (Hamel et al., 2015), but also suggests these viruses spread from the inoculation site and egress into the media. MAYV infection in human skin is not well characterized and to our knowledge, our data is the first to show MAYV infection in human skin ex vivo. Alphaviruses such as CHIKV also replicate in human skin cell populations (Matusali et al., 2019). Human skin fibroblasts, both primary and immortal, are susceptible to CHIKV in vitro (Ekchariyawat et al., 2015; Sourisseau et al., 2007; Wichit et al., 2017). CHIKV-infected human skin explants showed the presence of viral RNA at 24h and 48h post-infection when infected with a larger inoculum (10^5^) (Bryden et al., 2020). Interestingly, both primary human and immortalized keratinocytes poorly replicate CHIKV by restricting replication at a post-endosomal fusion step despite the high multiplicity of infection *in vitro* (MOI=50)(Bernard et al., 2015).

### MAYV and ZIKV induce inflammatory expression in the skin

Expression of pro-inflammatory signals from skin resident cell populations are critical for recruiting circulating myeloid cells to the site of infection(Pasparakis et al., 2014). Since this model utilizes skin detached from the microvasculature, one of the primary limitations is the influx of inflammatory cells to the site of infection. Therefore, we assessed the expression of inflammatory targets which recruit myeloid cell populations to the site of infection. Chemo-attractive chemokines CCL2, CCL3, CCL4, and CXCL8 were upregulated during MAYV and ZIKV infection (Figure 4). In other studies, DENV-infected keratinocytes express CCL2, CCL3, CCL4, CCL5, CXCL8 in a similar trend at 48h post-infection (Duangkhae et al., 2018). The same DENV-infected culture also shows the expression of IL-1α, IL-1β, IL-10, and CCL20 at the same timepoint (Duangkhae et al., 2018). WNV infection induces the expression of antiviral cytokines such as IFN-β, IL-28a, and IL-29 at 24h and 48h post-infection in primary keratinocytes (Garcia et al., 2018). Additionally, expression of CXCL1, CXCL2, CXCL8, and CCL20 and cytokines IL-6 and TNFα are also induced at the same time points (Garcia et al., 2018).

**Figure 4.**
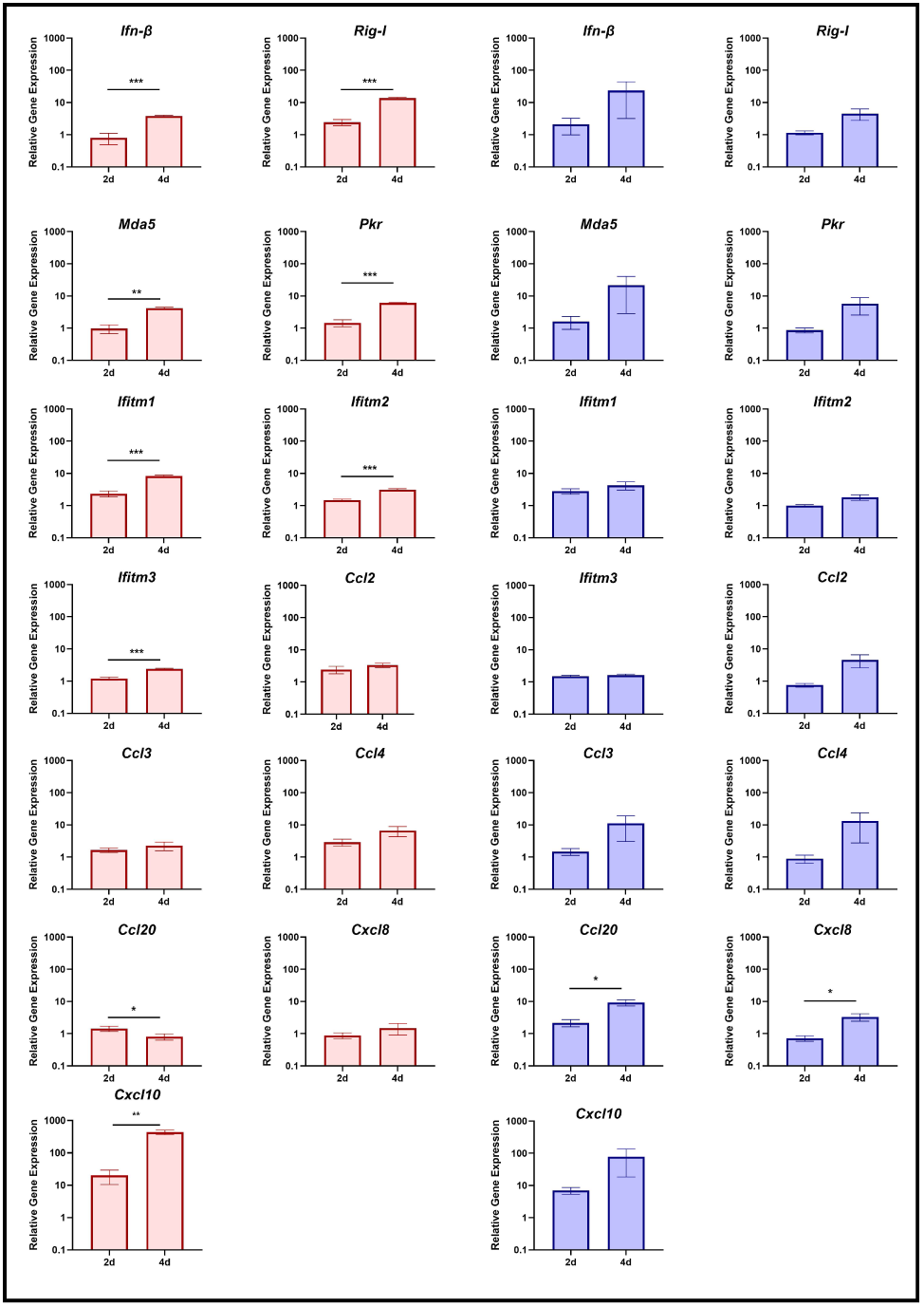
qPCR inflammatory gene expression of arbovirus infected human skin. qPCR analysis of MAYV infected tissues (red) and ZIKV infected tissues (blue) at 2 and 4 days post-infection. Data shown is the relative fold change (Livak method 2^-ΔΔCt^) of virus infected tissue compared to mock infected tissue, normalized to 18s ribosomal RNA. Expression levels from 2d to 4d were compared for statistical difference. Error bars indicate the standard error of the mean (SEM). Data is representative of experimental means with standard deviations of independent experiments with repeat using n=4 biological replicates per group. Student’s t test with Welch’s correction was used to determine statistical difference *p<0.05; **p<0.001; ***p<0.0001.

Elevated expression levels of non-TLR receptors of viral ligands, RIG-I, MDA5, and PKR were also found in ZIKV and MAYV infected tissues. RIG-I and MDA5 are cytosolic helicases that detect viral RNA and are critical for triggering innate immune responses to flaviviruses and alphaviruses (Akhrymuk et al., 2016; Chow et al., 2018; Kell & Gale, 2015). PKR is a pattern recognition receptor (PRR) and an IFN-inducible-gene product that triggers signaling cascades responsible for further type I IFN activation and inhibition of viral protein synthesis, among other functions (Munir & Berg, 2013). Both MAYV and ZIKV infection induced an increase in the expression of RIG-I, MDA5, and PKR in these tissues (Figure 4). This corresponds to previous findings of synthetic dsRNA poly(I:C) initiating antiviral signaling in keratinocytes (Kalali et al., 2008). The natural expression of pro-inflammatory and antiviral genes in skin explants provides a suitable model for studying human skin’s immediate immune signaling events.

Interferon-induced transmembrane proteins (IFITMs) are strongly upregulated by type I and type II interferon and localize to plasma and endocytic membranes to restrict virus entry (Bailey et al., 2014; Zhao et al., 2019). Given IFITM molecules show restriction of ZIKV (Savidis et al., 2016) and MAYV (Franz et al., 2021) infection, we assessed the expression of IFITM1, -2, and -3 in human skin tissue infected with either ZIKV or MAYV (Figure 4). IFITM1 levels were elevated during ZIKV infection as expected, however there was not a significant increase from 2 to 4 dpi. IFITM2 slightly increased above baseline at 4d post-infection, while IFITM3 remained near basal expression levels. MAYV infection induced a more robust IFITM expression. IFITM1 was elevated at 2 and 4 dpi with statistically higher expression at 4 dpi. IFITM2 and IFITM3 were slightly elevated at 2 dpi but significantly increased at 4 dpi. Further experimentation would be necessary to better understand the role of these antiviral molecules in the skin in relation to virus infection at the mosquito bite site.

## Conclusion

Skin is the largest organ of the human body that interfaces with external stimuli, including pathogens, and is permissive to arbovirus infection (Garcia et al., 2017). Blood-feeding mosquitoes deliver virus from infected salivary glands along with a cocktail of pharmacologically active molecules. Several studies show mosquito-derived salivary factors inducing greater morbidity and higher viremia when delivered with a pathogen in animal models (Pingen et al., 2016). This highlights the importance of understanding mosquito-specific factors active during transmission and the need to characterize such molecules in human skin. Human skin can be used as an *ex vivo* experimental site of infection for alpha- and flaviviruses. Both MAYV and ZIKV replicate in human skin to 4 days post-infection and shed into the skin culture media. Infection in the skin leads to gene expression of chemotactic factors responsible for recruiting inflammatory cells such as neutrophils and macrophages. In addition, innate antiviral responses are active and induced after viral infection. Histological analysis showed no major pathological changes; however, immunohistochemical analysis showed widespread MAYV and ZIKV infection in human skin. Mosquito-borne virus replication in human skin can be utilized to further investigate mosquito transmission to humans and the associated salivary mediators that work in concert to facilitate blood acquisition and pathogen delivery.

## Data Availability

All data produced in the present work are contained in the manuscript

## Abbreviations used

(ZIKV): Zika virus
(MAYV): Mayaro virus
(CHIKV): Chikungunya virus
(SPOV): Spondweni virus
(USUV): Usutu virus
(BSL-2): biological safety level 2
(IRB): Institutional Review Board
(HI-FBS): heat-inactivated fetal bovine serum
(qRT-PCR): quantitative reverse transcriptase PCR
(LAS-X): Leica Application Suite X
(MOI): multiplicity of infection
(PRR): pattern recognition receptor
(IFITMs): Interferon-induced transmembrane proteins
(SEM): standard error of the mean

## Acknowledgment

The authors would like to acknowledge the anonymous donors for providing skin tissue used in this study.

## Funding

The study described in this manuscript was funded by Departmental Start-up funds and SUNY Empire Innovation Professorship funds to ST

## Conflict of interest

The authors state no conflict of interests.

## Author contributions

Conception: ST and ATE

Experimental design: ATE, JF, MG, ST

Resources: ST, PU, JM

Acquisition of data and analysis: ATE, PU

Interpretation of data: ATE and ST

Supervision: ST

Writing-Original Draft preparation: ATE

Writing-Review and Editing: ATE, JF, MG, PU, ST

Funding acquistion: ST

## Notes

### Competing Interest Statement

The authors have declared no competing interest.

